# Effects of oral hygiene and food intake on tongue swab testing for tuberculosis in South Africa

**DOI:** 10.64898/2026.08.19.26360845

**Authors:** Angelique K. Luabeya, Alaina M. Olson, Danelle van As, Kate Hadley, Rachel C. Wood, Simba Mabwe, Christel Petersen, Alexander J. Yan, Kris M. Weigel, Paul Yager, Mark Hatherill, Gerard A. Cangelosi

## Abstract

The WHO has recommended tongue swabs (TS) as alternative samples for microbiological diagnosis of tuberculosis. We evaluated the effects of oral hygiene and food/drink intake on TS performance in South Africa. Food/drink intake prior to sampling marginally decreased *Mycobacterium tuberculosis* DNA signal strength, but neither behaviour decreased diagnostic sensitivity.

## Background

The World Health Organization recently recommended tongue swabs (TS) as alternative diagnostic samples when sputum cannot be collected for microbiological diagnosis of pulmonary tuberculosis (TB). TS sampling combined with molecular testing for *Mycobacterium tuberculosis* DNA has exhibited accuracy that meets or exceeds WHO target product profiles for non-sputum sample tests (1-7).

Swab samples collected from the tongue dorsum have shown better sensitivity than swab samples from other parts of the oral cavity (4), possibly because *Mycobacterium tuberculosis* (MTB) cells expelled during exhalation or coughing become entrained in bacterial biofilm on the tongue dorsum. Factors affecting the oral biofilm and the tongue epithelium, such as nutrition, disease, medication, or oral hygiene, might affect the accumulation or retention of MTB on the tongue dorsum. For example, comparisons of qPCR quantification cycle (Cq) values revealed greater DNA yields from TS collected early in the morning compared to later in the day (4). Morning behaviors such as eating and oral hygiene might affect the amount of MTB DNA detectable by TS. Because the TS method remains novel, there is a need to better understand the impacts of patient behaviors on its performance. Therefore, we evaluated the effects of oral hygiene (OH) and food/drink intake (FD) on the diagnostic performance of TS testing.

## Methods

The study was conducted in Worcester, Western Cape, South Africa. Enrollment of participants with confirmed TB at local health services clinics has been described elsewhere (2, 4). The population in the present study was the Cohort 1 described previously (2) .

Participants were included in the study if they gave written informed consent, were 18 years or older, had sputum GeneXpert Ultra results positive for MTB, and had been on TB treatment for less than 72 hours. The study focused on diagnostic sensitivity relative to sputum testing, and did not include participants who were not diagnosed with TB. The University of Cape Town and University of Washington ethics committees approved this study.

Each enrolled participant (N=100) with sputum GeneXpert-confirmed TB was asked to provide TS samples across three days. All sampling was completed within three days of initiation of treatment. The participants were trained to self-collect TS (8) and to follow oral hygiene (OH) and food or drink (FD) intake instructions before sample collection. OH behaviors included tooth brushing, dental flossing, tongue scraping, and use of mouth wash. Each participant collected TS under three different conditions of OH and FD. For the first condition (C1), patients took no food or drink, and performed no oral hygiene (OH-, FD-), before TS collection. For the second condition (C2), OH was allowed but there was no FD intake before TS collection (OH+, FD-). For the third condition (C3), there was no OH but FD intake was allowed before TS collection (OH-, FD+). We did not collect information on the type of food ingested nor the type of oral hygiene performed before sampling.

Participants were assigned to 3 groups. Each group started the first sample collection under either C1, C2, or C3 conditions, and rotated accordingly. This was done to minimize biases that might otherwise result from the order in which the conditions occurred. Ultimately, all participants provided TS under all three OH and FD intake conditions on different days. Baseline characteristics and behaviours (age, sex, BMI, ethnicity, level of education, smoking, alcohol usage, background oral hygiene habits and associated conditions, chronic diseases, medications, duration of TB symptoms before seeking care, area of residence, type of housing, and household size) were recorded. They did not differ significantly between the three groups **(Table 1)**.

**Table 1.** Baseline characteristics of participants enrolled in the study.

| Variables (N, %) | All participants (N=100) | Group 1 (N=34) | Group 2 (N=32) | Group 3 (N=34) |
| --- | --- | --- | --- | --- |
| Age (mean, SD) | 36.2 (11.3) | 36.8 (11.7) | 35.9 (10.6) | 35.7 (11.8) |
| Gender (male) % | 63 (63.0) | 24 (70.6) | 19 (59.4) | 20 (58.8) |
| Race (mixed race) | 77 (77.0) | 26 (76.5) | 26 (81.3) | 25 (73.5) |
| (black) | 23 (23.0) | 8 (23.5) | 6 (18.8) | 9 (26.5) |
| **Level of education** |  |  |  |  |
| No schooling | 2 (2.0) | 0 | 1 (3.1) | 1 (2.9) |
| Primary | 20 (20.0) | 7 (20.6) | 6 (18.8) | 7 (20.6) |
| Post primary | 78 (78.0) | 27 (79.4) | 26 (78.1) | 26 (76.5) |
| **Employment status** |  |  |  |  |
| Unemployed | 69 (69.0) | 24 (70.6) | 23 (71.9) | 22 (64.7) |
| Casual | 12 (12.0) | 2 (5.9) | 4 (12.5) | 6 (17.7) |
| Permanent | 19 (19.0) | 8 (23.5) | 5 (15.6) | 6 (17.7) |
| **Type of housing if known** |  |  |  |  |
| Farm | 5 (5.1) | 2 (5.9) | 1 (3.1) | 2 (6.1) |
| Flat | 12 (12.1) | 4 (11.8) | 3 (9.4) | 5 (15.2) |
| House | 50 (50.5) | 17 (50.0) | 15 (46.9) | 18 (54.6) |
| Informal settlement | 32 (32.3) | 11 (32.4) | 13 (40.6) | 8 (24.2) |
| **Smoking habits (Yes)** |  |  |  |  |
| Current | 69 (69.0) | 17 (50.0) | 26 (81.3) | 26 (76.5) |
| Former | 16 (16.0) | 10 (29.4) | 2 (6.3) | 4 (11.7) |
| Never | 15 (15.0) | 7 (20.6) | 4 (12.5) | 4 (11.8) |
| **Drinking alcohol (Yes)** |  |  |  |  |
| Current alcohol drinker | 35 (35.0) | 14 (41.2) | 9 (28.1) | 12 (35.3) |
| Former Alcohol drinker | 15 (15.0) | 6 (17.7) | 3 (9.4) | 6 (17.7) |
| Never used Alcohol | 50 (50.0) | 14 (41.2) | 20 (62.5) | 16 (47.1) |
| **Previous TB (Yes)** | 37 (37.0) | 17 (50.0) | 10 (31.3) | 10 (29.4) |
| **HIV (Yes)** | 35 (35.0) | 15 (44.1) | 8 (25.0) | 12 (35.3) |
| **Diabetes (Yes)** | 4 (4.0) | 2 (5.9) | 0 | 2 (5.9) |
| **Oral hygiene habits and associated conditions** |  |  |  |  |
| Brush teeth (Yes) | 92 (92.0) | 30 (88.2) | 31 (96.9) | 31 (91.2) |
| Dental floss (Yes) | 5 (5.0) | 3 (8.8) | 0 | 2 (5.9) |
| Clean tongue (Yes) | 92 (92.0) | 30 (88.2) | 31 (96.9) | 31 (91.2) |
| Brush tongue | 89 (89.0) | 28 (82.4) | 30 (93.8) | 31 (91.2) |
| Use mouth wash (Yes) | 2 (2.0) | 2 (5.9) | 0 | 0 |
| Gum bleeding (Yes) | 24 (24.0) | 8 (23.5) | 9 (28.1) | 8 (23.5) |
| Bad breath (Yes) | 20 (20.0) | 10 (29.4) | 5 (15.6) | 5 (14.7) |
| Dentist visits Yes) | 77 (77.0) | 22 (64.7) | 26 (81.3) | 29 (85.3) |

Frozen samples were transferred to the University of Washington, Seattle, WA, where they were processed and tested in blinded fashion by using a previously described manual qPCR method that employed sequence-specific magnetic separation (SSMaC) (9). This early version of the method detected a single MTB genetic element, IS*6110*, and may have been less sensitive than newer multi-target laboratory methods (10). It may also have been less sensitive than commercial, near-point of care (nPOC) methods that were recently endorsed by WHO (1). It was used because the study was completed before the newer methods became available. However, the methodology was uniform across all OH and FD conditions in this study.

One swab per condition per participant was processed and tested. A qPCR Cq threshold of ≤38 was used to define TS positivity, as described previously (4). Lower qPCR Cq values correspond with stronger TS signals. Samples that yielded Cq values >38 were defined as “TB negative” (4). The sample size was calculated to detect a Cq mean difference of 1.3 within each paired comparison (paired t-test, alpha 0.05, power 0.80, two-tailed). A Chi-square test was used to compare proportions. Statistical associations were tested using STATA version 12.1 (StataCorp LP,4905 Lakeway Drive, College Station, Texas 77845 USA).

## Results

Results from qPCR (either Cq values or “TB negative”) across all three treatment conditions were available for 93 of the 100 participants. Other participants were excluded because six participants’ swabs were not tested for one or more condition, and one participant’s TB status was not confirmed at the time of analysis. A negative TS result was defined as either: 1) A Cq value greater than the predefined cutoff threshold value (Cq>38), indicating equal to or less amplification than expected background signal (4); or 2) a test readout of “Undetermined”, indicating no qPCR amplification. Results from qPCR were false negative for 25 OH-FD-samples, 18 OH+FD-, and 23 OH-FD+ participants. All other TS samples yielded true-positive results. Percent sensitivities relative to sputum Xpert status were compared between the three sampling conditions. Small differences were observed, but no differences were statistically significant by a paired z-score test of proportions (significance level 0.05) (**Table 2, top)**.

**Table 2.** Percent sensitivities and mean Cq values of TS results under three sample collection conditions.

| Sampling conditions | TS percent sensitivity relative to sputum Xpert (n/N) |  |
| --- | --- | --- |
| OH-FD- | 73.1% (68/93) |  |
| OH+FD- | 80.6% (75/93) |  |
| OH-FD+ | 75.4% (70/93) |  |
| Comparisons of mean Cq values in positive samples (N comparisons) | Mean Cq values (SD) | P value (2-tailed paired t test) |
| OH-FD-/ OH+FD- (65) | 28.25 (4.5)/28.73 (5.7) | 0.45 |
| **OH-FD-/ OH-FD+ (62)** | **28.08 (4.2)/29.26 (4.9)** | **0.04** |
| OH+FD-/ OH-FD+ (66) | 28.8 (5.9)/29.1 (5.0) | 0.6 |

Among the TS samples that yielded positive qPCR results, mean Cq values were compared between the conditions. Three pairwise comparisons were possible **(Table 2, bottom)**. Two comparisons (OH-FD-vs. OH+FD-; and OH+FD-vs. OH-FD+) did not exhibit significant differences by paired t test (significance level 0.05). One comparison (OH-FD-vs OH-FD+) exhibited a marginally significant difference (*p* = 0.04), with slightly higher Cq values (weaker signal) in the FD+ condition. However, this did not translate to a detectable decrease in percent sensitivity relative to sputum Xpert-based diagnosis (**Table 2, top**).

## Discussion

We evaluated parameters that may impact the performance and robustness of TS as a novel sample type for TB diagnosis. In terms of percent sensitivity relative to sputum Xpert-based diagnosis, OH and FD intake before TS did not have detectable impacts on TS diagnostic accuracy. This is an encouraging outcome that may help facilitate the implementation of TS sampling.

When looking at quantitative signal strengths as indicated by qPCR Cq values among participants with positive TS, we found that FD intake prior to sampling were associated with weakened qPCR signals, as indicated by a higher mean Cq value. This difference was marginally significant by paired t test. FD residues may inhibit qPCR reactions or increase the non-target biomass on the tongue, potentially interfering with swab-based detection of MTB DNA. Such effects appear to be small given the lack of detectable impacts on percent sensitivity.

A limitation of this study is that it was completed prior to the introduction of more sensitive multi-target TS tests (1, 10). In addition to differences in genetic targets, our method used chemical conditions and enzymatic approaches that may differ from newer methods. The effects of assay inhibitors present in FD may differ significantly across these platforms. Moreover, the study did not include symptomatic people without TB, so the effects of FD and OH on test specificity were not determined. However, our study provides a framework by which widely-used, WHO-recommended TS test platforms can, in the future, be assessed for impacts of patient behaviours and other variable parameters.

An additional limitation is that we did not collect chest X-rays, sputum culture results, or sputum Xpert Ultra semi-quantitative data to evaluate the severity of the disease. These data would have strengthened interpretation of the findings and allowed for a more nuanced analysis taking disease severity into account.

A structural limitation was that OH, FD, and other behaviours were self-reported and could not be verified. Inaccurate self-reporting could have obscured associations. Notably, we have observed that TS donors in another study can sometimes be self-conscious about their OH (11). The overall sample size of N = 93 participants with qPCR results may have been insufficient to detect differences that might be detectable with a larger sample size.

Finally, because the study was conducted in a single geographical location, generalizability may be limited, especially given regional differences in eating and drinking habits and cuisines.

Despite these limitations, to our knowledge, this study is the first systematic attempt to assess the impacts of certain common behaviours on TS results. Marginally significant impacts were identified on the level of qPCR signal strength (Cq value), but not on the level of diagnostic accuracy (percent sensitivity). This is an encouraging finding as the TS method progresses toward implementation.

## Data Availability

All data produced in the present study are available upon reasonable request to the authors

## Acknowledgements

This work was supported by the Gates Foundation (INV-004527) and NIAID grants R01AI139254 and R21AI185407. The authors are indebted to study staff and participants. We thank Copan Diagnostics for providing swabs.

